# Estimating the health impact of vaccination against 10 pathogens in 98 low and middle income countries from 2000 to 2030

**DOI:** 10.1101/19004358

**Authors:** Xiang Li, Christinah Mukandavire, Zulma M Cucunubá, Kaja Abbas, Hannah E Clapham, Mark Jit, Hope L Johnson, Timos Papadopoulos, Emilia Vynnycky, Marc Brisson, Emily D Carter, Andrew Clark, Margaret J de Villiers, Kirsten Eilertson, Matthew J Ferrari, Ivane Gamkrelidze, Katy Gaythorpe, Nicholas C Grassly, Timothy B Hallett, Michael L Jackson, Kévin Jean, Andromachi Karachaliou, Petra Klepac, Justin Lessler, Xi Li, Sean M Moore, Shevanthi Nayagam, Duy Manh Nguyen, Homie Razavi, Devin Razavi-Shearer, Stephen Resch, Colin Sanderson, Steven Sweet, Stephen Sy, Yvonne Tam, Hira Tanvir, Quan Minh Tran, Caroline L Trotter, Shaun Truelove, Kevin van Zandvoort, Stéphane Verguet, Neff Walker, Amy Winter, Neil M Ferguson, Tini Garske (Vaccine Impact Modelling Consortium)

## Abstract

**Background:** The last two decades have seen substantial expansion of childhood vaccination programmes in low and middle income countries (LMICs). Here we quantify the health impact of these programmes by estimating the deaths and disability-adjusted life years (DALYs) averted by vaccination with ten antigens in 98 LMICs between 2000 and 2030.

**Methods:** Independent research groups provided model-based disease burden estimates under a range of vaccination coverage scenarios for ten pathogens: hepatitis B (HepB), *Haemophilus influenzae* type b (Hib), human papillomavirus (HPV), Japanese encephalitis (JE), measles, *Neisseria meningitidis* serogroup A (MenA), *Streptococcus pneumoniae*, rotavirus, rubella, yellow fever. Using standardized demographic data and vaccine coverage estimates for routine and supplementary immunization activities, the impact of vaccination programmes on deaths and DALYs was determined by comparing model estimates from the no vaccination counterfactual scenario with those from a default coverage scenario. We present results in two forms: deaths/DALYs averted in a particular calendar year, and in a particular annual birth cohort.

**Findings:** We estimate that vaccination will have averted 69 (2.5-97.5% quantile range 52-88) million deaths between 2000 and 2030 across the 98 countries and ten pathogens considered, 35 (29-45) million of these between 2000-2018. From 2000-2018, this represents a 44% (36-57%) reduction in deaths due to the ten pathogens relative to the no vaccination counterfactual. Most (96% (93-97%)) of this impact is in under-five age mortality, notably from measles. Over the lifetime of birth cohorts born between 2000 and 2030, we predict that 122 (96-147) million deaths will be averted by vaccination, of which 58 (39-75) and 38 (26-52) million are due to measles and Hepatitis B vaccination, respectively. We estimate that recent increases in vaccine coverage and introductions of additional vaccines will result in a 72% (61-79%) reduction in lifetime mortality caused by these 10 pathogens in the 2018 birth cohort.

**Interpretation:** Increases in vaccine coverage and the introduction of new vaccines into LMICs over the last two decades have had a major impact in reducing mortality. These public health gains are predicted to increase in coming decades if progress in increasing coverage is sustained.

## Introduction

Vaccines have been responsible for substantial reductions in mortality ^1–5^ and vaccination is among the most cost-effective health interventions ^6,7^. In addition to direct protection afforded to vaccinated individuals, high levels of vaccination coverage offer indirect protection (herd immunity) to the remaining unvaccinated individuals in a population. Impact is seen on a timescale which is vaccine-dependent: for some childhood diseases (such as measles, rotavirus, pneumococcal disease), impact is seen rapidly, while for human papillomavirus (HPV) and hepatitis B, impacts are commonly seen over a much longer timescale in the reduction of adult morbidity and mortality.

The World Health Organization (WHO) introduced the Expanded Programme on Immunization (EPI) in 1974 ^8^. The programme - supported by UNICEF and global donors - succeeded in delivering substantial increases in coverage of routine childhood vaccines; for instance, global coverage of DTP3 increased from just over 20% in 1980 to over 75% in 1990 ^9^. However, as coverage plateaued in the 1990s, concerns grew around the sustainability of these gains, eventually leading to the formation of Gavi (the Vaccine Alliance) in 1999 ^10^. Gavi’s mission is to sustain and increase coverage and provide improved access to new vaccines in low and middle income countries (LMICs) ^11^. Since its founding, it has supported immunisation of more than 700 million children in LMICs ^12^. Global targets for vaccination have also continued to grow in ambition: the Global Vaccine Action Plan (GVAP) framework was launched in 2012 by WHO with the aim of preventing millions of deaths by 2020 through access to vaccines in all countries. This was further reinforced by target 3.8 of the Sustainable Development Goals calling for access to “vaccines for all” by 2030 ^13^.

Due to the limitations in completeness and quality of death registration and disease surveillance systems in many LMICs, it is not possible in most of the world to directly measure the impact of vaccination programmes on mortality and morbidity. Mathematical models are therefore an essential tool for extrapolating from available disease burden and pathogen surveillance data to generate impact estimates. Modelling is also required to generate projections of the impact of future vaccine coverage to inform investment planning.

To improve the quality and coordination of vaccine impact assessment, the Vaccine Impact Modelling Consortium (VIMC) was formed in late 2016 with the support of Gavi and the Bill & Melinda Gates Foundation. VIMC currently comprises 18 modelling groups coordinated by a secretariat at Imperial College London, and it models vaccination impact against diseases caused by ten different pathogens across 98 countries (about 69% of the world’s population in 2018), including the 73 countries currently eligible for Gavi support.

This paper presents the first complete set of vaccine impact estimates generated since VIMC was formed, quantifying impact over annual birth cohorts between 2000-2030 in terms of deaths and disability adjusted life-years (DALYs) averted. The ten pathogens included were hepatitis B (HepB), *Haemophilus influenzae* type B (Hib), human papillomavirus (HPV), Japanese encephalitis (JE), measles, *Neisseria meningitidis* serogroup A (MenA), *Streptococcus pneumoniae* (prevented by the pneumococcal conjugate vaccine - PCV), rotavirus (rota), rubella virus and yellow fever virus (YF). VIMC does not currently assess the impacts of diphtheria, tetanus and pertussis (DTP) vaccine ^1^, cholera vaccine and polio vaccines. VIMC’s vaccine impact estimates are used to support the monitoring of existing vaccination programmes and inform future investment strategy.

## Methods

Modelling groups from different institutions with disease-specific expertise provided the pathogen-specific vaccine impact estimates (Table 1). A total of 20 pathogen-specific mathematical models were used by VIMC to produce the vaccine impact estimates presented here: two models for each pathogen other than HepB, which had three, and yellow fever, which had one. Including multiple models for each pathogen enables some assessment of the impact of structural uncertainty in models. Each model represents the impact of vaccine coverage and efficacy on national level disease burden (and in some cases disease transmission dynamics) to estimate vaccine impact. Model descriptions and the list of 98 LMICs included in the analysis are available in the supplementary information (SI).

**Table 1:**
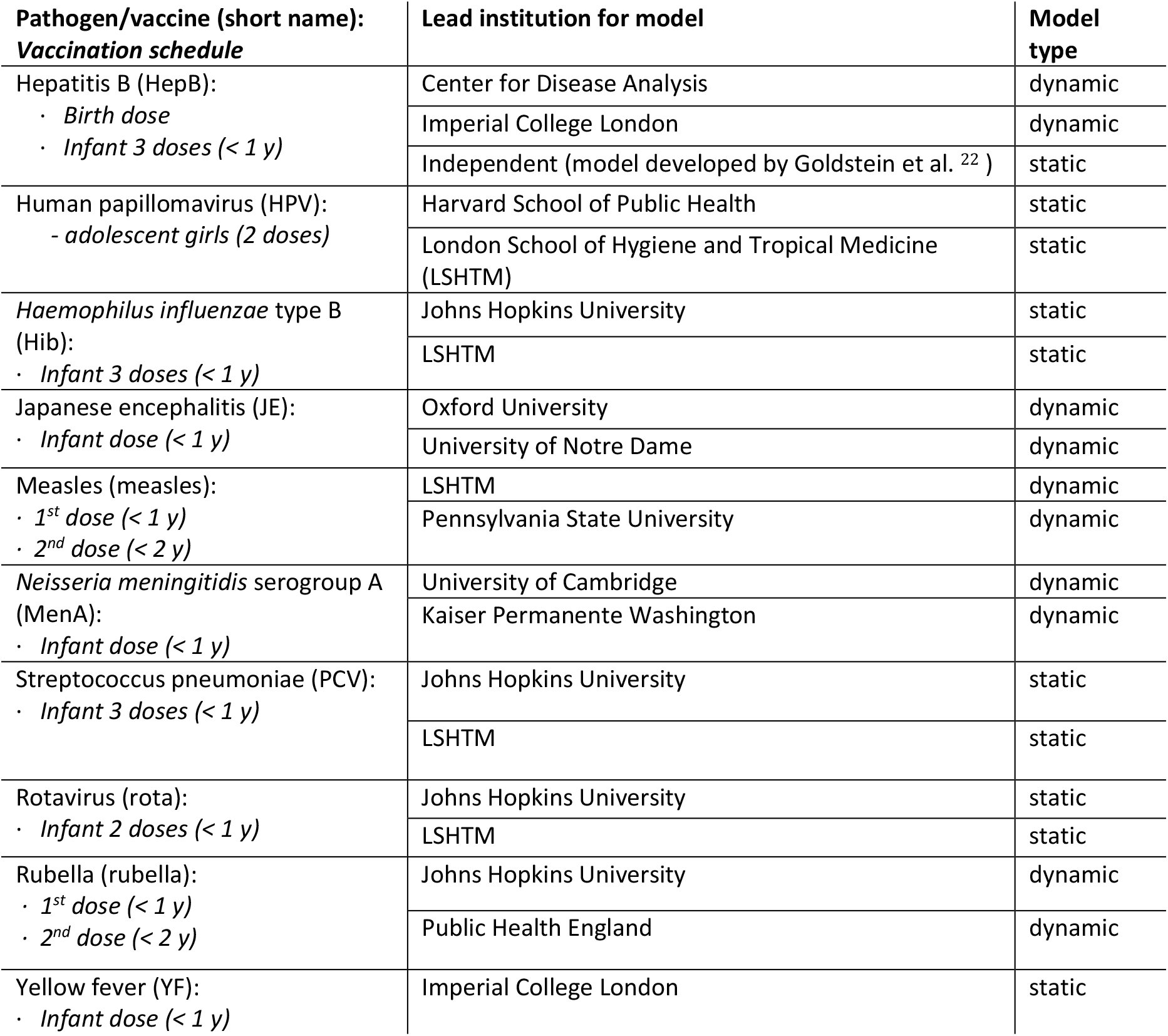
Pathogen names, vaccination schedules, modelling groups and model types included in VIMC. Static models only model the direct effect of vaccination on vaccinated cohorts assuming that pathogen transmission intensity is not modified by vaccination coverage. Dynamic models simulate infectious disease transmission dynamics and model both the direct effect of vaccination on vaccinated cohorts and indirect/herd effect of vaccination on unvaccinated populations. See SI model descriptions for more details and citations for each model.

| Pathogen/vaccine (short name):<br>*Vaccination schedule* | Lead institution for model | Model type |
| --- | --- | --- |
| Hepatitis B (HepB):<br>· *Birth dose*<br>· *Infant 3 doses (< 1 y)* | Center for Disease Analysis | dynamic |
|  | Imperial College London | dynamic |
|  | Independent (model developed by Goldstein et al. <sup>22</sup>) | static |
| Human papillomavirus (HPV):<br>- *adolescent girls (2 doses)* | Harvard School of Public Health | static |
|  | London School of Hygiene and Tropical Medicine (LSHTM) | static |
| *Haemophilus influenzae* type B (Hib):<br>· *Infant 3 doses (< 1 y)* | Johns Hopkins University | static |
|  | LSHTM | static |
| Japanese encephalitis (JE):<br>· *Infant dose (< 1 y)* | Oxford University | dynamic |
|  | University of Notre Dame | dynamic |
| Measles (measles):<br>· *1<sup>st</sup> dose (< 1 y)*<br>· *2<sup>nd</sup> dose (< 2 y)* | LSHTM | dynamic |
|  | Pennsylvania State University | dynamic |
| *Neisseria meningitidis* serogroup A (MenA):<br>· *Infant dose (< 1 y)* | University of Cambridge | dynamic |
|  | Kaiser Permanente Washington | dynamic |
| Streptococcus pneumoniae (PCV):<br>· *Infant 3 doses (< 1 y)* | Johns Hopkins University | static |
|  | LSHTM | static |
| Rotavirus (rota):<br>· *Infant 2 doses (< 1 y)* | Johns Hopkins University | static |
|  | LSHTM | static |
| Rubella (rubella):<br>· *1<sup>st</sup> dose (< 1 y)*<br>· *2<sup>nd</sup> dose (< 2 y)* | Johns Hopkins University | dynamic |
|  | Public Health England | dynamic |
| Yellow fever (YF):<br>· *Infant dose (< 1 y)* | Imperial College London | static |

Standardised demographic data (live births per year, death rates) based on the United Nations World Population Prospects (UNPD World Population Prospects - UNWPP 2017 ^14^) was used for all 98 countries. Similarly, standardised national level estimates of vaccination coverage for each vaccine considered were provided by the VIMC secretariat to each group. Past coverage in all countries for 1980-2016 was obtained from WHO/UNICEF Estimates of National Immunization Coverage (WUENIC) as published in July 2017 ^15^, whose online coverage estimates are updated annually. Future coverage estimates from 2017 to 2030 were based on Gavi’s strategic demand forecast as of October 2017, for the countries eligible for Gavi support. Gavi’s operational forecasts assume likely dates of vaccine introduction based on non-binding expressions of interest from eligible countries, applications to Gavi for vaccine support, intended introductions as reported to the World Health Organization (WHO), and assessment of country capacity to introduce a specific vaccine in a specific time frame. Following introduction, coverage of new vaccines are typically assumed to reach coverage of a reference vaccine (e.g., DTP3) within two to three years, after which coverage is assumed to increase 1 percent per year up until a maximum of 90% or 95% depending on the vaccine ^1^. For the 25 countries considered not supported by Gavi, and for years after 2030 for countries in Gavi’s portfolio, an annual 1% increase in coverage was assumed from 2017 up to a maximum of 90% or the historic high coverage achieved (if >90%). For newly introduced vaccines with only an introduction date and no Gavi coverage forecast, we assumed that coverage would increase to the same coverage as pentavalent (HepB-Hib-DTP3) vaccine in that country in the first three years and subsequently increase by 1% per year. Estimates of numbers of vaccines received per child by year were generated from these coverage estimates and projections assuming independence of coverage between vaccines.

Disease burden was quantified by deaths and DALYs. DALYs measure the years of healthy life lost due to premature death and disability from the disease, and are the sum of years of life lost (YLLs) through premature mortality and years lived with disability (YLDs). Estimates of deaths and DALYs were stratified by age. No discounting or weighting was applied in the calculation of DALYs.

Age-stratified pathogen-specific disease burden estimates (deaths and DALYs) for annual birth cohorts between 2000 to 2030 were generated by each modelling group. Corresponding estimates for the counterfactual scenario, assuming no vaccination had occurred after 2000, were also produced. Supplementary immunisation activities (SIAs), other immunisation campaigns and second doses were modelled when relevant.

For rubella, only disease burden from congenital rubella syndrome (CRS) ^16^ was assessed, since rubella only causes mild disease in infected persons. Since the occurrence of CRS in infants is determined by the vaccination and infection status of the mother while pregnant, disease burden for CRS was reported by the age of the mother (at the time of birth) and not to that of the infant.

The impact of vaccination was assessed by comparing the counterfactual (‘no vaccination’) scenario with the vaccination scenarios. Two forms of aggregation were used to present the results: by calendar year and by year of birth. The former assesses the difference in burden between the vaccination and no-vaccination scenarios for a specific year, and gives a cross-sectional view of impact. The latter sums disease burden across every year of life for each yearly birth cohort (born between 2000 and 2030) before again calculating the difference between vaccination and no-vaccination scenarios, and therefore gives a lifetime view of vaccine impact.

The population-attributable benefit of vaccination for each pathogen was estimated by calculating the proportion of annual deaths due to each pathogen that would be prevented by vaccination.

We use model averaging to derive impact estimates, with each model for a pathogen given equal weighting. Uncertainty in model estimates was assessed by generating 200 sets of estimates from each model, probabilistically sampling over model parameter uncertainty. The same randomly sampled sets of parameters were used for both vaccination and no-vaccination model runs, allowing uncertainty in vaccine impact to be assessed. Central pathogen-specific estimates presented here represent averages over all such samples from every model available for each pathogen. The 95% (2.5 and 97.5% quantiles) credible intervals presented for each pathogen are derived by combining the probabilistic distributions of estimated impact from all the models available for that pathogen.

Estimates involving aggregation across pathogens were generated via a bootstrap approach under the simplifying assumption that the drivers of uncertainty in each model are independent of those in any other model. For each model, a random sample of the statistic of interest was drawn from the 200 probabilistic runs. These model-specific samples were combined by averaging them across models of the same pathogen and then summing the resulting pathogen-specific estimates across pathogens. Means and 2.5 and 97.5% quantiles of 100,000 such bootstrap samples were calculated to derive central estimates and 95% credible intervals.

The main text of this paper focuses on presenting vaccine impacts on mortality; more detailed estimates of mortality impacts and estimates of DALYs averted by vaccination are given in the SI.

## Results

The average number of vaccines received per child increased consistently and substantially across the great majority of the 98 countries (Table S1) between 2000 and 2018 (Figure 1A-C). Both increases in coverage of existing vaccines (e.g. measles-containing vaccines) and the introduction of new vaccines (e.g. rotavirus) contribute to this overall trend (Figure 1D). Routine vaccination against JE and rotavirus began to be introduced from 2006, PCV in 2010, HPV in 2014 and MenA in 2016. In addition, coverage increases likely reflect the increase in countries eligible for Gavi support (Table S2).

**Figure 1.**
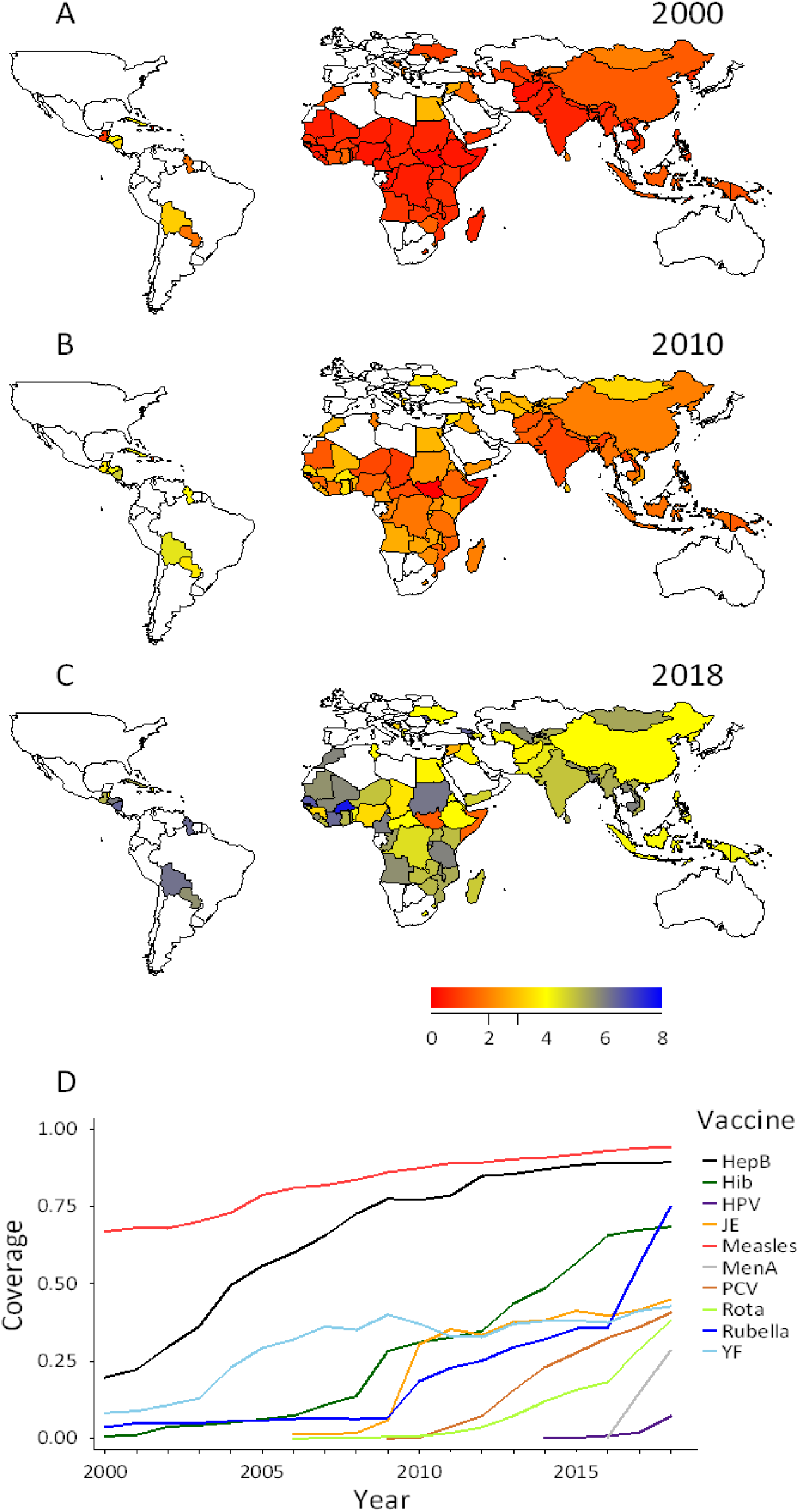
Map of vaccine coverage across the 10 pathogens considered, calculated as mean number of vaccines received per child in (A) 2000, (B) 2010, (C) 2018. The colour scale shows the expected number of vaccines received per child in each country. (D) Routine vaccine coverage for each pathogen, from 2000 to 2018, averaged across all 98 countries except for JE, MenA and YF which were averaged across the 16, 26 and 32 endemic countries for those pathogens, respectively. The average was obtained by dividing total vaccine doses by the total eligible population.

To assess the impact of vaccine use on mortality, we quantify the expected burden of disease in the counterfactual scenario of no vaccination (Figure 2, red curves) and estimate the impact of observed and projected vaccination coverage on that baseline mortality (Figure 2, blue curves). Depending on the pathogen considered, long-term trends in disease prevalence interact with global population growth and aging to result in a variety of projected trends in disease-specific mortality in the counterfactual no-vaccination scenario. However, for all but two pathogens, vaccination between 2000 and 2030 is estimated to cause substantial reductions in the mortality burden in the same time period. The two exceptions are HepB and HPV where most mortality in that time window is due to infections that have already occurred, due to the typically long time delay between infection and severe outcomes for those infections. While HepB vaccination coverage is now relatively high and projected to increase further, most of the impacts of this will only be seen after 2030. For HPV, most countries have yet to introduce vaccination.

**Figure 2:**
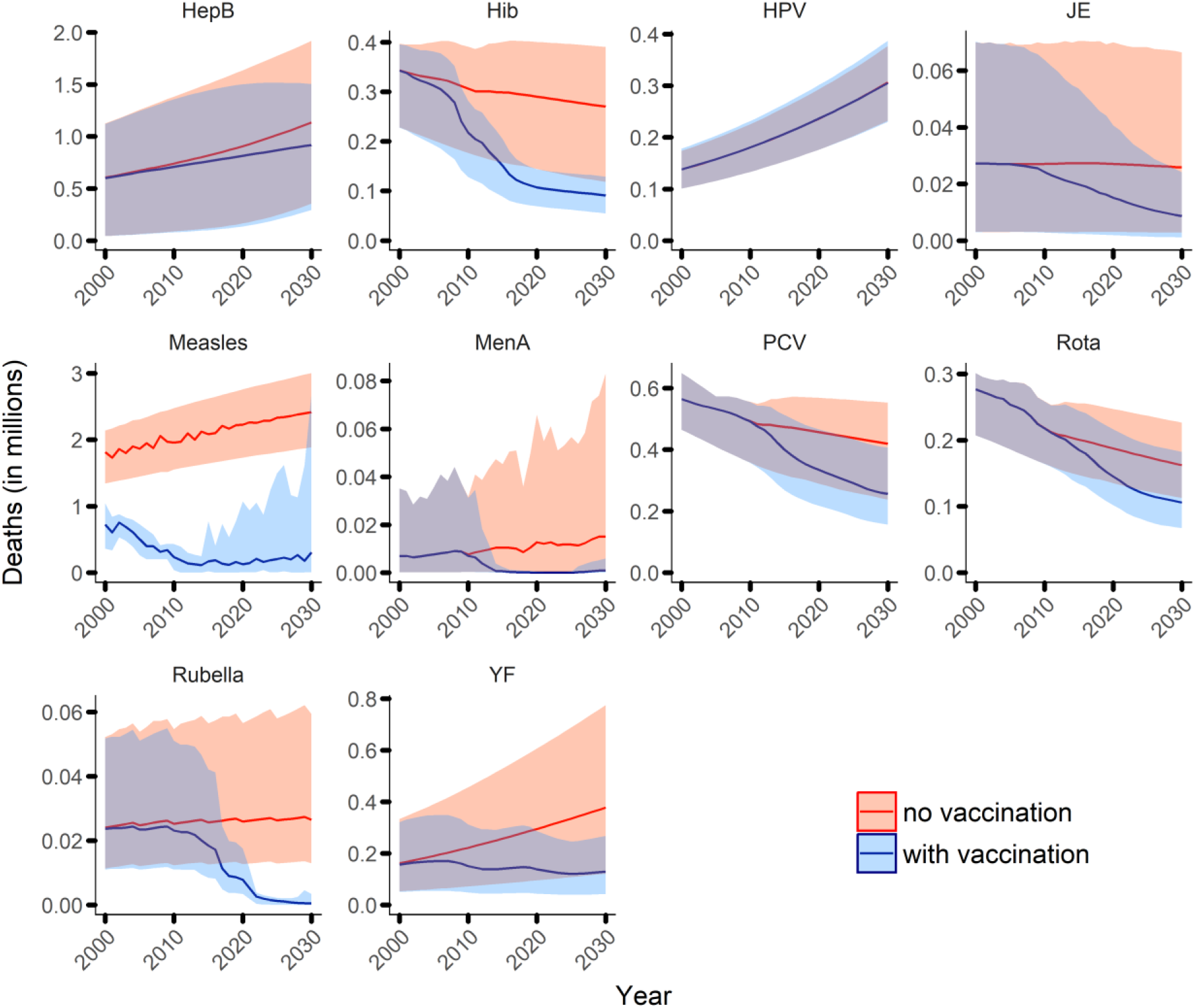
Estimates of disease-specific deaths by calendar year from 2000 to 2030 across all 98 countries, for default projections of vaccine coverage and counterfactual (no vaccination) coverage. Continuous blue and red lines show estimates of deaths for default and no vaccination coverage scenarios for all ages, respectively. The shaded areas show the 95% credible region.

The age distribution of mortality varies dramatically across the 10 pathogens as a result of differences in their epidemiology: HepB- and HPV-attributable mortality primarily affects those over 40 years of age, YF, MenA and JE are epidemic diseases which largely affect those under 30 (due to natural immunity acquired with age in older adults), while mortality for all other pathogens is nearly entirely focused in the under-5s (though note that mortality due to infection with rubella is attributed to the age of the mother and is therefore focused in women of childbearing age).

Figure 3 presents the estimated numbers of deaths averted by vaccination in the 98 countries (see also SI Tables 5(a-h)). Two views are given: total deaths averted by calendar year (Figure 3A), and deaths averted over the lifetime of each annual birth cohort (Figure 3B).

**Figure 3:**
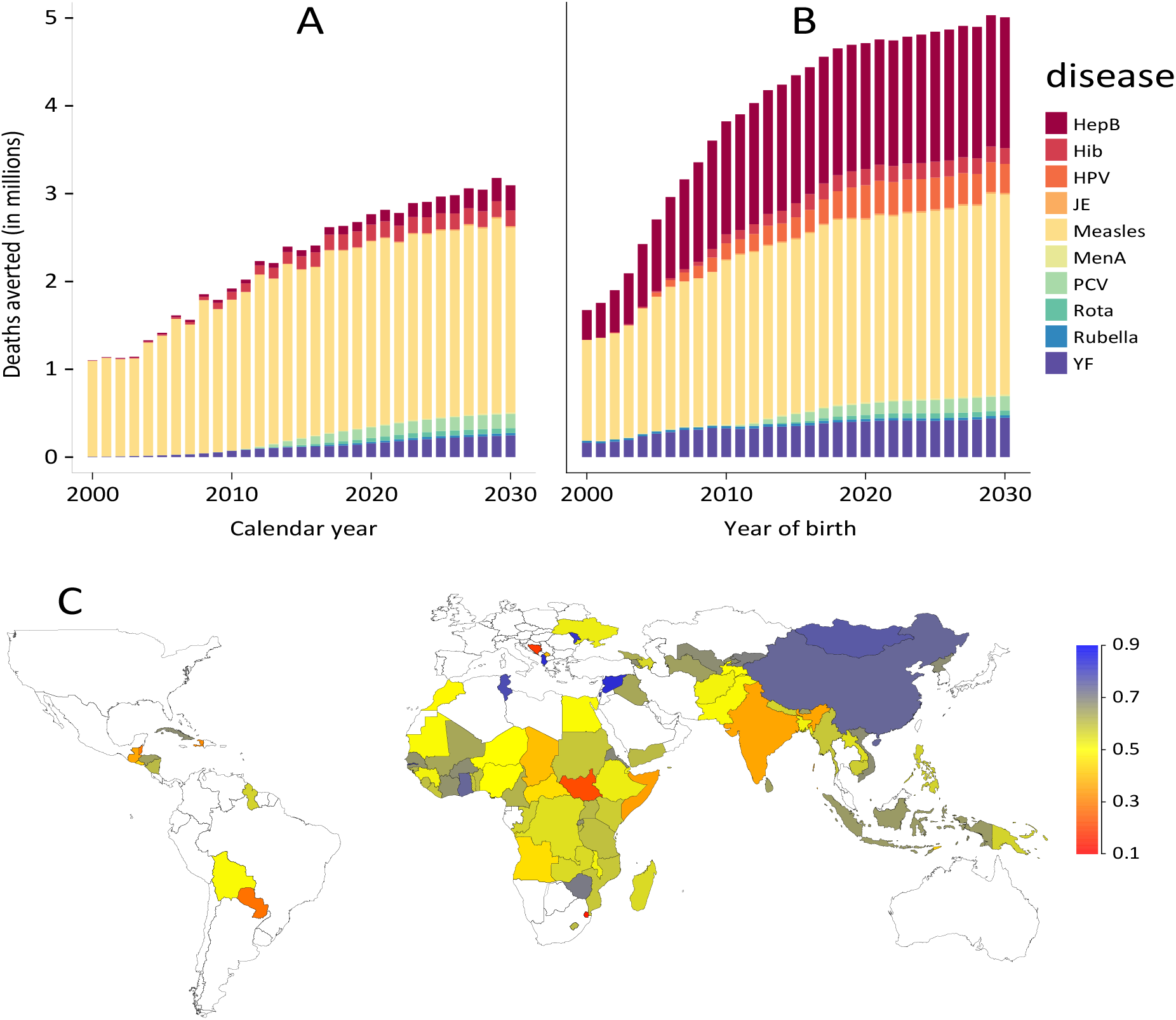
Central estimates of deaths averted in the 98 countries: A) by calendar year (summing across all ages) and pathogen; B) by year of birth (summing across lifetime) and pathogen; C) proportion of lifetime deaths due to the 10 pathogens in the no-vaccination counterfactual that are predicted to be averted by vaccination, by country across 2000-2018 birth cohorts.

Looking by calendar year, we estimate a total of 69 (52-88) million deaths averted between 2000 and 2030, 35 (29-45) million of which were averted between 2000 and 2018. In the 73 Gavi countries, the corresponding values are 66 (49-82) million deaths averted between 2000-2030, 33 (27-42) million of which were averted between 2000-2018.

Measles vaccination has the largest impact, with 56 (39-74) million deaths averted (54 (38-70) million in the 73 Gavi countries).

Considering deaths averted by birth cohort (Figure 3B and Table 2), the longer-term impact of increasing coverage of HepB vaccination becomes clearer. However, since the great majority of HepB deaths (due to liver disease) occur in those over 45 years of age, the impact of vaccination in the 2000-2030 time period will only really start to be seen from 2040 onwards. Similar arguments apply to HPV vaccination (though coverage is substantially lower than for HepB): cervical cancer deaths largely occur in women over 50, leading to a delay of nearly 40 years between vaccination and its direct impact on mortality. Thus, summing vaccination impact over the full lifetimes of the 2000-2030 birth cohorts gives a total of 122 (96-147) million deaths averted in total, 65 (47-69) million of which are in the under-5s. In the 73 Gavi countries, the corresponding values are 106 (81-130) million deaths averted in the 2000-2030 birth cohorts, 65 (49-83) million of which are in the under-5s. Table 2 summarises deaths averted by pathogen and illustrates that in the 2000-2030 birth cohorts over 53% (44-63%) of deaths averted are in children under 5 years.

**Table 2:** Estimated total deaths averted (in thousands) by vaccination and deaths averted per thousand individuals vaccinated in different annual birth cohort ranges across the 98 countries considered, stratified by pathogen. Both lifetime and under-5 deaths averted are shown.

| Vaccine | Birth cohorts | Deaths averted (1000s) lifetime | Deaths averted per 1000 vaccinated individuals, lifetime | deaths averted (1000s), <5 | Deaths averted per 1000 vaccinated individuals, <5 |
| --- | --- | --- | --- | --- | --- |
| HepB | 2000-2018 | 20000 (14000-30000) | 13 (8.9-19) | 280 (58-1100) | 0.2 (0-0.6) |
|  | 2019-2030 | 18000 (11000-27000) | 9.9 (6.4-15) | 250 (66-890) | 0.1 (0-0.5) |
| Hib | 2000-2018 | 1600 (690-2300) | 2.8 (1.3-4.2) | 1600 (690-2300) | 2.8 (1.3-4.2) |
|  | 2019-2030 | 2200 (800-3500) | 2.4 (0.9-3.7) | 2200 (800-3500) | 2.4 (0.9-3.7) |
| HPV | 2000-2018 | 2600 (1600-3300) | 15 (10-20) | 0 (0-0) | 0 (0-0) |
|  | 2019-2030 | 4300 (3800-4800) | 12 (10-14) | 0 (0-0) | 0 (0-0) |
| JE | 2000-2018 | 170 (17-420) | 0.4 (0-1) | 57 (4-150) | 0.1 (0-0.3) |
|  | 2019-2030 | 230 (25-600) | 0.5 (0.1-1.2) | 96 (9-250) | 0.2 (0-0.5) |
| Measles | 2000-2018 | 32000 (24000-42000) | 7.2 (5.5-9.3) | 32000 (24000-42000) | 7 (5.5-9.2) |
|  | 2019-2030 | 26000 (11000-34000) | 9.2 (4.2-12) | 25000 (12000-33000) | 9 (4.5-12) |
| MenA | 2000-2018 | 220 (110-380) | 0.9 (0.5-1.5) | 22 (1-60) | 0.1 (0-0.2) |
|  | 2019-2030 | 200 (83-360) | 0.8 (0.3-1.3) | 36 (2-96) | 0.1 (0-0.4) |
| PCV | 2000-2018 | 590 (300-1100) | 3 (1.5-5.1) | 590 (300-1100) | 3 (1.5-5.1) |
|  | 2019-2030 | 1800 (790-3300) | 2.4 (1.1-4.3) | 1800 (790-3300) | 2.4 (1.1-4.3) |
| Rota | 2000-2018 | 130 (84-190) | 1 (0.6-1.4) | 130 (84-190) | 1 (0.6-1.4) |
|  | 2019-2030 | 640 (400-900) | 0.8 (0.5-1.1) | 640 (400-900) | 0.8 (0.5-1.1) |
| Rubella | 2000-2018 | 510 (250-1200) | 0.4 (0.2-0.9) | 0 (0-0) | 0 (0-0) |
|  | 2019-2030 | 340 (160-770) | 0.1 (0.1-0.3) | 0 (0-0) | 0 (0-0) |
| YF | 2000-2018 | 5600 (1800-12000) | 16 (5-32) | 550 (170-1200) | 1.5 (0.5-3.2) |
|  | 2019-2030 | 5000 (1600-11000) | 18 (5.9-37) | 570 (170-1300) | 2.1 (0.6-4.3) |
| Total | 2000-2018 | 64000 (52000-78000) | 6.8 (5.5-8.2) | 35000 (27000-45000) | 3.7 (3-4.7) |
|  | 2019-2030 | 58000 (42000-72000) | 5.4 (3.9-6.6) | 31000 (17000-39000) | 2.8 (1.6-3.6) |
|  | 2000-2030 | 120000 (95000-150000) | 6 (4.7-7.2) | 65000 (48000-84000) | 3.2 (2.3-4.1) |

The extent to which vaccination reduces overall mortality due to the 10 pathogens varies substantially by country (Figure 3C), largely due to historical variations in vaccination coverage, but also due to variation in the epidemiology of some pathogens by country. Summing across the 98 countries we consider, we estimate vaccination will prevent 72% (61-79%) of the mortality associated with these 10 pathogens in the 2018 annual birth cohort. Considering only under-5 mortality, this proportion rises to 76% (59-83%). In the 73 Gavi countries, the corresponding values are a 71% (60-78%) reduction in all-age mortality, and a 77% (60-83%) reduction in under-5 mortality.

In the period 2000-2018, we estimate that vaccination in the 98 countries reduced overall mortality and under-5 mortality due to these pathogens by 44% (36-57%) and 57% (52-64%), respectively. In the 73 Gavi countries, these reductions were 47% (39-58%) and 57% (52-64%), respectively. For the period 2019-2030, we project these reductions will respectively rise to 60% (33-74%) and 77% (42-85%) in all 98 countries and 64% (35-76%) and 77% (42-85%) in the 73 Gavi countries.

It is informative to place these estimates in their demographic context. Taking the 2018 birth cohort as an example, and using UNWPP demographic estimates (see Methods), we estimate that under-5 mortality in the 98 countries would be 45% (33-56%) higher in the absence of any vaccination against the ten pathogens. The total number of deaths (occurring at any age) averted in the 2018 birth cohort represent 4.2% (3.3-5.1%) of the live births making up that cohort.

Total impact reflects vaccination coverage as well as underlying disease burden and vaccine effectiveness. We therefore also examine the relative impact of each vaccine, by quantifying deaths averted per vaccinated individual for each pathogen (Table 2 and SI Tables 6(a-h)). This highlights that while measles and PCV have the largest relative impact on under-5 mortality, vaccines against HPV, HepB and YF have the largest impact per person vaccinated. YF and HPV vaccination have the largest relative impact of all the vaccines considered, with central estimates for both of over 16 (5-32) deaths averted per 1000 persons vaccinated for the 2000-2018 birth cohorts.

Since most of the pathogens considered (namely Hib, JE, measles, PCV, rota, rubella) largely result in under-5 mortality, the impact of vaccination on DALYs largely mirrors the impact on mortality (Fig S3). Since HepB and HPV related deaths are focused in the over 50s, mortality contributes fewer years of life lost (YLLs) for these pathogens, but morbidity contributes higher years lived with disability for both infections (HepB particularly). Estimates of vaccine impact in DALYs averted are presented in the SI.

## Discussion

This study represents the largest scale study of health impacts of immunisation in LMICs yet undertaken, covering vaccination programs against 10 pathogens and evaluating impact in 98 countries. It represents an advance on past work ^1^ in its scale (in both antigens and countries considered) and in the emphasis of VIMC on standardising model inputs (vaccine coverage and demography) and outputs (age-specific mortality and DALYs by year of birth cohort and age). Such standardisation allows impacts to be combined across and compared between vaccines.

We find that immunisation programmes in the 98 countries considered will result in individuals born in 2018 experiencing 72% (61-79%) lower mortality due to those 10 pathogens over their lifetime than they would otherwise. Furthermore, in the absence of vaccination, we estimate that all-cause under-5 mortality would be 45% (33-56%) higher than currently observed. These impacts are a testament to both the public health benefit of vaccines overall and the sustained investment in increasing global vaccination coverage in the last two decades. They also highlight what might be lost if current vaccination programmes are not sustained, and thus provide quantitative evidence supporting both donor and country investments in vaccination programmes.

Deriving these impact estimates is far from straightforward: cause-specific mortality data in the LMICs considered is very limited, making direct observational assessment of impact challenging. However, to inform monitoring and decision-making, countries and international organizations such as Gavi require projections of potential impact under a range of investment scenarios. To fill these gaps, mathematical and statistical models can be used to extrapolate data on levels of current infection (such as case detection from active surveillance) and/or past infection (such as serosurveys) to sites and countries without such data. They can also be used to project future trends given information about vaccine coverage.

In addition, our study has focused on quantifying uncertainty in vaccine impact. Given the limited explicit data on pathogen-specific disease burden available in many of the 98 countries considered, nearly all models need to extrapolate from settings where data are available to those where data are absent. This, together with imperfect knowledge of aspects of the epidemiology of each pathogen (e.g. case fatality ratios, transmissibility, disease progression rates) means that uncertainty in vaccine impact estimates from a single model can be substantial. Here this uncertainty is quantified probabilistically, with each modelling group providing 200 models runs spanning the range of parametric uncertainty in their models.

A second source of uncertainty is structural: different modelling groups make different subjective choices about how to represent disease epidemiology and may use different data for model parameterisation. In addition, the models within VIMC vary substantially: in their type (static cohort models versus transmission-dynamic models), in their complexity (e.g. in the representation of age effects), and in their approaches to calibration and validation (from formal statistical likelihood approaches to more ad-hoc calibration). VIMC therefore includes at least two models for each pathogen (with the exception of YF) and combines results from different models to derive central estimates of impact and to better quantify underlying uncertainty (see Methods).

A limitation of our current analysis is that we do not currently evaluate uncertainty in demographic estimates and estimates of past and future vaccine coverage. Developing principled approaches to doing so is a topic of current research, but is made challenging by the limited information available on uncertainty in UNWPP demographic estimates and in WHO UENIC and Gavi operational forecast vaccine coverage estimates.

The majority of models within VIMC adopt a ‘bottom-up’ approach to modelling disease burden and thus the impact of vaccination. These models represent time- and age-varying pathogen-specific infection or disease rates in each country, then model mortality as affecting a fraction of those infected by applying a case fatality ratio (generally estimated from a combination of longitudinal epidemiological studies and surveillance data) to resulting case incidence estimates. As the disease burden attributed to each pathogen is modelled separately, there is a theoretical risk of overestimating deaths due to a failure to account for competing causes of mortality, particularly in the under-5s, where most mortality is concentrated. However, in the 2018 annual birth cohort, UNWPP projections estimate all-cause under-5 mortality at 5% for the 98 countries considered. We estimate the 10 pathogens we consider will cause approximately one seventh of this (0.69% (0.5-1.2%)). With such low absolute proportions, the effect of competing hazards of death on overall mortality estimates is negligible. Conversely, when assessing deaths averted, we consider the counterfactual of no vaccination for each vaccine antigen separately, subsequently summing across all vaccines, since one child’s life can be saved multiple times.

For most pathogens, we currently model infection risk as homogeneous within individual countries (the exceptions being the YF and JE models). Furthermore, no models in this study account for geographic or socioeconomic clustering of vaccine coverage, or for any potential correlation between access to healthcare (including vaccines) and disease risk. Thus, we may be ignoring disadvantaged sub-populations in countries with lower than average access to vaccines and/or higher than average intrinsic exposure to infection. Subnational stratification of vaccine impact estimates are a priority for future work but require similarly fine-grained estimates of vaccine coverage ^17^ and disease burden.

Last, in making long-term projections of disease burden and intervention impact, it is necessary to make assumptions about the likely improvements in treatment and disease outcomes in future decades. This is a particular issue for HepB and HPV, where cancer screening and treatment services can make a substantial difference to disease-related mortality ^18^, or for measles where decreasing background under-five mortality can significantly reduce case-fatality ratios ^19^. The HPV and HepB models included in VIMC currently make conservative (i.e. relatively pessimistic) assumptions about improvements in cancer screening and treatment in low income countries.

More generally, the estimates provided here should not be viewed as immutable; our understanding of the epidemiology and disease burden caused by all 10 pathogens continues to improve, and models of those diseases should likewise continue to be refined. In addition, future vaccine coverage is unlikely to precisely match the coverage projections used here. Thus, the outputs - whether estimates of the impact of past immunisation activities or projections of future impact - will also change. However, the results in this paper provide the most comprehensive and definitive assessment to date of the impact of the dramatic advances in immunisation coverage in LMICs in the last two decades.

Finally, our analysis highlights where the greatest gains from future investments in improving vaccine coverage are to be made. We predict increasing HPV coverage in girls will avert more deaths per person vaccinated than any other immunisation activity, while increasing PCV coverage will give the largest reductions in under-5 mortality. However, it is even more important not to allow the coverage gains achieved since 2000 to slip back. This requires continued political commitment, funding, civil society engagement (in promoting benefits and countering hesitancy), improving public trust and confidence in the safety and efficacy of vaccines ^20^, and strengthening immunisation programmes through education, training and supervision ^21^.

## Data Availability

Please refer to the tables in the supplementary information.

## Acknowledgements

We thank Gavi, the Vaccine Alliance and the Bill & Melinda Gates Foundation for funding VIMC (BMGF grant number: OPP1157270). We also acknowledge joint Centre funding from the UK Medical Research Council and Department for International Development, which supported aspects of VIMC’s work.

## References

1 Lee LA, Franzel L, Atwell J, et al. The estimated mortality impact of vaccinations forecast to be administered during 2011-2020 in 73 countries supported by the GAVI Alliance. Vaccine 2013; 31 Suppl 2: B61–72.

2 Whitney CG, Zhou F, Singleton J, Schuchat A, Centers for Disease Control and Prevention (CDC). Benefits from immunization during the vaccines for children program era - United States, 1994-2013. MMWR Morb Mortal Wkly Rep 2014; 63: 352–355.

3 Roush SW, Murphy TV, Vaccine-Preventable Disease Table Working Group. Historical comparisons of morbidity and mortality for vaccine-preventable diseases in the United States. JAMA 2007; 298: 2155–2163.

4 Andre FE, Booy R, Bock HL, et al. Vaccination greatly reduces disease, disability, death and inequity worldwide. Bull World Health Organ 2008; 86: 140–146.

5 van Wijhe M, McDonald SA, de Melker HE, Postma MJ, Wallinga J. Effect of vaccination programmes on mortality burden among children and young adults in the Netherlands during the 20th century: a historical analysis. Lancet Infect Dis 2016; 16: 592–598.

6 (55) Ozawa S, Mirelman A, Stack ML, Walker DG, Levine OS. Cost-effectiveness and economic benefits of vaccines in low- and middle-income countries: a systematic review. Vaccine 2012; 31: 96–108.

7 Horton S. Cost-Effectiveness Analysis. In: Disease Control Priorities:Improving Health andReducing Poverty. 2017: 147–156.

8 Keja K, Chan C, Hayden G, Henderson RH. Expanded programme on immunization. World Health Stat Q 1988; 41: 59–63.

9 Wallace AS, Ryman TK, Dietz V. Overview of global, regional, and national routine vaccination coverage trends and growth patterns from 1980 to 2009: implications for vaccine–preventable disease eradication and elimination initiatives. J Infect Dis 2014; 210 Suppl 1: S514–22.

10 Wittet S. Introducing GAVI and the global fund for children’s vaccines. Vaccine 2000; 19: 385–386.

11 Lob-Levyt J. Contribution of the GAVI Alliance to improving health and reducing poverty. Philos Trans R Soc Lond B, Biol Sci 2011; 366: 2743–2747.

12 Gavi: Mid-Term Review report 2016-2020. Gavi: https://www.gavi.org/library/publications/gavi/gavi-2016-2020-mid-term-review-report/, 2018 https://www.gavi.org/library/publications/gavi/gavi-2016-2020-mid-term-review-report/ (accessed Aug 21, 2019).

13 U N. Transforming our world: The 2030 agenda for sustainable development. A/RES/70/1. 2015; published online Oct 21. https://sustainabledevelopment.un.org/post2015/transformingourworld/publication (accessed June 3, 2019).

14 UNWPP. The 2017 Revision of the United Nations World Population Prospects. 2017. https://esa.un.org/unpd/wpp/. (accessed Jan 21, 2018).

15 WHO. Immunization monitoring. 2017. https://apps.who.int/immunization_monitoring/globalsummary/timeseries/tswucoveragedtp3.h tml (accessed July 18, 2017).

16 Duszak RS. Congenital rubella syndrome--major review. Optometry 2009; 80: 36–43.

17 Mosser JF, Gagne-Maynard W, Rao PC, et al. Mapping diphtheria-pertussis-tetanus vaccine coverage in Africa, 2000-2016: a spatial and temporal modelling study. Lancet 2019; 393: 1843–1855.

18 Torre LA, Siegel RL, Ward EM, Jemal A. Global Cancer Incidence and Mortality Rates and Trends-- An Update. Cancer Epidemiol Biomarkers Prev 2016; 25: 16–27.

19 Portnoy A, Jit M, Ferrari M, Hanson M, Brenzel L, Verguet S. Estimates of case-fatality ratios of measles in low-income and middle-income countries: a systematic review and modelling analysis. Lancet Glob Health 2019; 7: e472–e481.

20 Larson HJ, Clarke RM, Jarrett C, et al. Measuring trust in vaccination: A systematic review. Hum Vaccin Immunother 2018; 14: 1599–1609.

21 WHO. Global Vaccine Action Plan 2011-2020. 2012. https://www.who.int/immunization/global_vaccine_action_plan/GVAP_doc_2011_2020/en/ (accessed Aug 6, 2019).

22 Goldstein ST, Zhou F, Hadler SC, Bell BP, Mast EE, Margolis HS. A mathematical model to estimate global hepatitis B disease burden and vaccination impact. Int J Epidemiol 2005; 34: 1329–1339.

